# Sex and APOE ε4 allele dose relate to longitudinal brain aging by amyloid status

**DOI:** 10.64898/2026.07.13.26357678

**Authors:** Reza Rajabli, Mahdie Soltaninejad, Sylvia Villeneuve, D. Louis Collins, the Alzheimer’s Disease Neuroimaging Initiative

## Abstract

**INTRODUCTION:** Sex and APOE ε4 influence Alzheimer’s disease risk, but whether they are associated with longitudinal structural brain aging and whether these associations depend on amyloid status is unclear. We tested these relationships using brain age gap (BAG), an MRI-derived marker of age-disproportionate brain atrophy.

**METHODS:** We trained a deep-learning brain-age model on 26,512 neurologically healthy UK Biobank participants and adapted it in 3,332 amyloid-negative cognitively normal MCSA and OASIS-3 scans. Without ADNI training, the model was applied to 4,399 T1-weighted MRI visits from 1,337 ADNI participants. Linear mixed-effects and survival models tested associations among sex, APOE ε4 allele dose, amyloid status, BAG, cognition, atrophy, and amyloid conversion; equivalence tests evaluated null effects.

**RESULTS:** Among amyloid-positive participants, women and men had similar baseline BAG, but BAG increased faster in women. APOE ε4 was associated with faster BAG increase in a dose-dependent manner. Higher and increasing BAG tracked hippocampal, fusiform, and whole-brain tissue loss in both amyloid groups. In amyloid-negative participants, BAG change was unrelated to four of five cognitive measures, with effects equivalent to zero within ±0.1 SD, whereas within-person BAG–cognition associations were evident across all five measures in amyloid-positive participants. Higher baseline BAG predicted progression from MCI to dementia (HR 1.67) and from cognitively normal to impaired (HR 1.40).

**DISCUSSION:** Sex and APOE ε4 allele dose were associated with longitudinal brain aging primarily among amyloid-positive participants. BAG tracked structural neurodegeneration and showed stronger cognitive associations among amyloid-positive participants, supporting its interpretation as a stage-dependent marker of Alzheimer-related neurodegeneration. Independent replication is needed.

## Background

Structural brain aging varies substantially across individuals, even at the same chronological age. One way to quantify this variability is to use machine-learning models that predict brain age from MRI. Comparing predicted brain age with chronological age yields the brain age gap (BAG), an estimate of age-disproportionate structural brain aging. Higher BAG values (i.e., older-appearing brains) have been associated with vascular risk factors^1^, neuropsychiatric disorders and poorer cognitive performance^2^, and cerebrovascular burden in the Alzheimer’s disease (AD) continuum^3^.

BAG has therefore been proposed as a marker of accelerated structural brain aging in AD, and amyloid and p-tau have been associated with greater BAG, although these associations may differ by disease stage^4^. An important unresolved question is whether BAG carries the same biological and clinical meaning across stages of the AD continuum. Structural brain aging reflects multiple biological processes, many of which are not specific to AD, and BAG cannot distinguish among them. Consequently, a similar BAG may carry different information in amyloid-negative and amyloid-positive individuals.

Sex is an important source of heterogeneity in AD. Women account for about two-thirds of AD dementia cases^5^, and longitudinal analyses have reported faster atrophy in women across stages from healthy aging through AD dementia, although in cognitively normal individuals differences may be confined to medial temporal regions^6^. However, findings from brain-age studies are inconsistent. Taylor et al.^7^ found faster increases in BAG in women in ADNI in contrasts involving progressive mild cognitive impairment, while Cumplido-Mayoral et al.^3^ reported higher BAG in cognitively unimpaired amyloid-positive tau-positive (A+T+) women, although without a significant sex-by-biomarker interaction. In contrast, Millar et al.^8^ found BAG about three years higher in men than women in a cohort studied for autosomal-dominant AD, with a cross-sectional offset also present in non-carriers. Two Baltimore Longitudinal Study of Aging analyses have reported greater male vulnerability to neurodegeneration, demonstrating steeper regional volume loss^9^ and steeper declines in brain volume after estimated amyloid onset^10^ in men. Ravndal et al.^11^ found faster decline in men in cognitively healthy adults. Similarly, a meta-analysis of approximately 58,000 individuals did not support a uniformly stronger APOE ε4 association with risk of AD in women^12^. These cohorts differ in composition and position along the AD continuum. If sex-related differences in BAG depend on amyloid status, variation in disease stage across cohorts could contribute to these apparently conflicting findings.

APOE ε4 is the strongest common genetic risk factor for late-onset AD. Whether APOE ε4 shows an allele dose-dependent association with longitudinal BAG, and to what extent this association is related to amyloid burden, remains unclear.

Estimating BAG in older clinical cohorts is also challenging because brain-age models may generalize poorly across datasets and age ranges owing to differences in age distributions and MR acquisition protocols between the training datasets, and the cohorts BAG is applied to^13^. Direct transfer can produce substantial domain shift, whereas naive fine-tuning may reduce previously learned representations^14^. We therefore used continual learning to adapt a pretrained model^13^ to intermediate datasets closer to the target population while keeping the analysis cohort out of training. We call the resulting model SPARROW.

In this study, we investigated how sex, APOE ε4 allele dose, and amyloid status are associated with longitudinal changes in BAG. Our primary analyses tested whether women show a faster increase in BAG than men, whether APOE ε4 is associated with BAG progression in an allele dose-dependent manner, and whether these associations differ according to amyloid status. We further examined, across the range of amyloid burden, when sex differences in the rate of BAG increase become apparent. Secondary analyses characterized the anatomical correlates of BAG and tested whether its associations with cognition differ between amyloid-negative and amyloid-positive individuals, including whether higher baseline BAG is associated with subsequent cognitive decline. The brain-age model was developed and adapted without training or fine-tuning on ADNI, allowing these associations to be evaluated independently.

## Methods

### Study Design and Data

We used a three-stage pipeline to develop and evaluate the SPARROW brain-age model while keeping the Alzheimer’s Disease Neuroimaging Initiative (ADNI) analysis cohort independent of model training. First, the base model was trained on 26,512 neurologically healthy participants from the UK Biobank (UKBB)^15^, following the general brain-age modeling framework described previously^13^. Participants with neurological, cerebrovascular, structural brain, or other conditions likely to alter brain structure were excluded; detailed exclusion criteria are provided in the Supplementary Methods.

Second, to extend the model to older ages and more heterogeneous imaging environments, we adapted it using 3,332 T1-weighted MRI scans from 1,777 cognitively unimpaired, amyloid-negative participants from the Mayo Clinic Study of Aging (MCSA)^16^ and Open Access Series of Imaging Studies (OASIS-3)^17,18^. Finally, model performance was evaluated in 262 cognitively unimpaired, amyloid-negative ADNI participants with MRI acquired within 45 days of the baseline cognitive exam. The SPARROW model was applied, without any further training or fine-tuning, to the full ADNI analysis cohort. This design allowed longitudinal associations with BAG to be evaluated without (re)training the brain-age model on the cohort used for the primary analyses. Cohort characteristics are summarized in Table 1.

**Table 1.** Datasets used for training, adaptation, testing and final analysis.

| Dataset | Subset | Purpose | Size (Subjects / Scans) | Sex (F / M / Unidentified) | Age at Baseline ([min, max], mean ± std) |
| --- | --- | --- | --- | --- | --- |
| UK Biobank (UKBB) | Neurologically Healthy | Training the Base Model | 26,512 / 26,512 | 13,693 / 12,819 / 0 | [44, 82], 63.29 ± 7.55 |
| Mayo Clinic Study of Aging (MCSA) | Amyloid-negative Healthy Controls | Fine-tuning the Base Model | 1,123 / 1,297 | 518 / 605 / 0 | [49.00, 89.40], 69.10 ± 10.22 |
| Open Access Series of Imaging Studies (OASIS-3) | Amyloid-negative Healthy Controls | Fine-tuning the Base Model | 654 / 2,035 | 298 / 199 / 157 | [42.69, 91.61], 66.69 ± 9.12 |
| Alzheimer’s Disease Neuroimaging Initiative (ADNI) | Amyloid-negative Healthy Controls | External Test | 262 / 262 | 144 / 118 / 0 | [55.1, 89.6], 71.91 ± 6.11 |
| | Full | Final Analysis | 1,337 / 4,399 | 644 / 693 / 0 | [55.0, 91.4], 72.75 ± 7.22 |

### Imaging Cohorts

#### UK Biobank (UKBB)

The UK Biobank (UKBB) is a large population-based resource designed to investigate determinants of health and disease using imaging, genetic, and other phenotypic data^15^. We used T1-weighted MRI acquired on 3T Siemens Skyra scanners using a standardized 3D MPRAGE protocol^19,20^. After applying the neurological and structural exclusion criteria described above, 26,512 participants were included for training the base brain-age model. Detailed acquisition parameters are provided in the Supplementary Methods.

#### Mayo Clinic Study of Aging (MCSA)

The Mayo Clinic Study of Aging (MCSA) is a longitudinal population-based cohort of older adults from Olmsted County, Minnesota, designed to investigate aging and cognitive decline^16^. We included 1,297 T1-weighted MRI scans from 1,123 cognitively unimpaired, amyloid-negative participants. Imaging was performed on 3T GE and Siemens Prisma scanners using 3D T1-weighted sequences. Amyloid status was determined from the PET component of the cohort. MCSA was used, together with OASIS-3, to adapt the pretrained brain-age model to older ages and imaging environments that differed from those represented in UKBB.

#### Open Access Series of Imaging Studies (OASIS-3)

OASIS-3 is an open-access longitudinal neuroimaging resource containing data collected across adulthood and older age^17,18^. We included 2,035 T1-weighted MRI scans from 654 cognitively normal, amyloid-negative participants. Scans were acquired on 3T Siemens systems using a 3D MPRAGE sequence, and amyloid status was determined from available PET data. OASIS-3 was combined with MCSA for model adaptation before application to ADNI.

#### Alzheimer’s Disease Neuroimaging Initiative (ADNI)

The Alzheimer’s Disease Neuroimaging Initiative (ADNI) is a multisite longitudinal study designed to characterize imaging, clinical, and biomarker changes across the Alzheimer’s disease continuum. We included 3D T1-weighted MRI from participants enrolled across ADNI-1, ADNI-Go, ADNI-2, ADNI-3, and ADNI-4. Imaging was acquired across multiple sites using 1.5T and 3T scanners from GE, Siemens, and Philips with standardized 3D MPRAGE or vendor-equivalent protocols. No ADNI data were used to train or adapt the brain-age model. ADNI therefore served as the independent cohort for model evaluation and for all primary longitudinal analyses. Detailed acquisition parameters for all cohorts are provided in the Supplementary Methods.

### Brain age model development and adaptation

SPARROW (Simple Pseudo-fully-convolutional Architecture with Residuals, Regularized, Optimized and Weight-consolidated) was based on the SFCN-reg architecture used in our previous work^13^ (the residual-block schematic is shown in Supplementary Figure S1). The model incorporates residual connections, Group Normalization, stochastic-depth and dropout regularization, and exponential moving averaging of model weights. Three independently initialized models were trained and their predictions averaged at inference.

To adapt the pretrained model to the older age distribution represented in MCSA and OASIS-3 while limiting loss of information learned from UKBB, we used a continual-learning procedure consisting of initial linear probing followed by low-learning-rate fine-tuning with Elastic Weight Consolidation^21^. Sampling and model-selection procedures accounted for repeated scans and the relative scarcity of participants at the oldest ages. Full architectural specifications, training hyperparameters, regularization procedures, and adaptation methods are provided in the Supplementary Methods.

### Brain Age Gap Estimation on ADNI

After adaptation, SPARROW was applied without further training or fine-tuning to all eligible ADNI T1-weighted MRI scans. No ADNI images were used to train or adapt the model. Brain age gap (BAG) was calculated at each visit as predicted brain age minus chronological age; positive BAG values therefore indicate an older-appearing brain relative to chronological age.

### Analysis sample and variables

The analysis included ADNI participants with sufficient biomarker information to classify them as amyloid-positive or amyloid-negative; participants without an amyloid classification were excluded. Of 4,534 MRI visits from 1,352 eligible participants, 18 failed imaging quality control and did not yield a BAG estimate, and 117 lacked intracranial volume (ICV), leaving 4,399 visits from 1,337 participants. Participants contributed a mean of 3.3 MRI visits (range, 1–14).

Time was defined as years from each participant’s baseline examination, with baseline set to zero. Demographic, clinical, cognitive, genetic, and biomarker variables were obtained from ADNIMERGE^22^. Cognitive measures included the 13-item Alzheimer’s Disease Assessment Scale–Cognitive Subscale (ADAS-Cog-13)^23^, Mini-Mental State Examination (MMSE), Clinical Dementia Rating Sum of Boxes (CDR-SB), Rey Auditory Verbal Learning Test (RAVLT) immediate recall, and Montreal Cognitive Assessment (MoCA).

BAG, baseline age, ADAS-Cog-13, education, and ICV were standardized using the baseline distribution of cognitively unimpaired, amyloid-negative participants (n = 256, comprising the 262 ADNI test-set participants after excluding five with missing ICV and one without a BAG estimate), ensuring that follow-up observations did not contribute to the reference distribution. One standard deviation of BAG in this reference group corresponded to 3.64 years. Visits missing variables required for a given analysis were excluded rather than imputed.

Regional brain volumes were obtained from FreeSurfer measures available through ADNIMERGE. Analyses of regional volumes included ICV as a covariate rather than normalizing regional volume by dividing by ICV^24^.

APOE ε4 was analyzed primarily as allele dose (0, 1, or 2 ε4 alleles). Amyloid status was analyzed both categorically and, where specified, using continuous measures of amyloid burden.

For continuous AV45 analyses, we used each participant’s first available nonmissing florbetapir PET SUVR in ADNIMERGE as a time-invariant measure of amyloid burden. These values were recorded at the study baseline exam for 728 of 910 participants (80%) and at follow-up exams for 182 (20%). AV45 values were standardized across participants in each fitted sample.

### Statistical Analysis

Longitudinal BAG trajectories were analyzed using linear mixed-effects models with participant-specific random intercepts and random slopes for time. Fixed effects in the primary model included sex, baseline age, education, intracranial volume (ICV), time from baseline, the education × time and ICV × time interactions, and the sex × time interaction. MRI field strength was excluded from the primary model because it was correlated with other covariates. Models were fitted by maximum likelihood using statsmodels version 0.15.0^25^.

Categorical predictors were entered using centered contrasts and continuous predictors were centered so that main effects represented differences at the average covariate profile. Sex was coded so that the sex coefficient represented the male-minus-female difference; consequently, a negative sex × time coefficient indicated a faster increase in BAG among women. Detailed contrast coding and centering procedures are provided in the Supplementary Methods.

When BAG was used as a longitudinal predictor, between-person and within-person BAG effects were separated^26^. When effects were presented separately for amyloid-positive and amyloid-negative participants, heterogeneity between groups was tested using interaction terms in the full sample rather than inferred from statistical significance within individual subgroups.

#### Primary analyses: sex, APOE ε4 allele dose, and amyloid status

The first primary analysis tested whether longitudinal BAG trajectories differed according to sex and amyloid status. This was evaluated using the sex × amyloid status × time interaction. Participants who were amyloid-negative at baseline but subsequently became amyloid-positive were excluded from the persistently amyloid-negative group. Analyses were repeated using continuous measures of amyloid burden to determine whether sex differences in BAG trajectories varied across the amyloid distribution.

For analyses using first available AV45 measurements, the Johnson–Neyman procedure^27,28^ was used to identify the range of amyloid burden over which the estimated sex difference in longitudinal BAG change was statistically distinguishable from zero. Results were expressed in the original amyloid measurement units and relative to the sample distribution.

The second primary analysis examined APOE ε4 using allele-dose categories (0, 1, or 2 alleles) to test whether it was associated with BAG level or longitudinal BAG change in a dose-dependent manner. We also tested whether these associations differed according to amyloid status.

#### Secondary analyses: characterization and clinical relevance of BAG

To characterize the structural correlates of BAG, we examined associations with hippocampal, entorhinal, middle temporal, fusiform, whole-brain, and ventricular volumes. Models included age, sex, education, ICV, and time. BAG-by-sex and BAG-by-amyloid interactions tested whether these structural associations differed between groups. False-discovery-rate correction was applied within each family of interaction tests.

We next examined whether the cognitive correlates of BAG differed according to amyloid status. For these models, each cognitive outcome was oriented so that higher scores indicated greater impairment and standardized across its available visits in the full analysis cohort. BAG was separated into participants’ mean BAG and visit-specific deviations from that mean. Associations were tested with ADAS-Cog-13, MMSE, CDR-SB, RAVLT immediate recall, and MoCA, adjusting for age, sex, education, ICV, and time. Models were evaluated separately within amyloid-positive and persistently amyloid-negative groups, with between-group differences assessed using interaction terms in the full sample. BAG-by-sex interactions were additionally examined to determine whether BAG–cognition associations differed between women and men.

Associations of BAG with AD-related biomarkers were examined using continuous florbetapir (AV45) PET SUVR, CSF Aβ42, and CSF p-tau181. The first available nonmissing value of each biomarker was selected separately; measurements were not required to be from the same visit. These analyses tested biomarker associations with BAG level and longitudinal change separately from the regional-volume analyses.

Among participants who were amyloid-negative at baseline, Cox proportional-hazards models^29^ tested whether baseline BAG was associated with subsequent conversion to amyloid positivity, adjusting for age, education, ICV, sex, and the number of amyloid assessments. APOE ε4 was included as a positive control.

Clinical progression was analyzed using Cox models, with baseline BAG and diagnosis defined at the first retained MRI scan. Follow-up ran from that scan to the first subsequent clinical visit showing progression from CN to MCI or dementia, or from MCI to dementia; participants without an event were censored at their last clinical visit with an available diagnosis. BAG models adjusted for baseline age, education, ICV, sex, APOE ε4 carrier status, and amyloid status; amyloid-stratified models omitted amyloid status. Models testing sex as the exposure adjusted for baseline age, education, ICV, and APOE ε4 carrier status.

For analyses in which evidence for a negligible association was substantively important, equivalence testing was performed using prespecified bounds of ±0.10 SD.

#### Sensitivity analyses

We assessed the robustness of the sex findings by adding, one at a time, baseline diagnosis × time, baseline ADAS-Cog-13 × time, APOE ε4 allele dose × time, MRI field strength × time, or age × time to the model and by removing ICV (Table 2). We also repeated the continuous AV45 analysis using only measurements recorded at the study baseline exam, retaining the same covariates and otherwise eligible longitudinal MRI visits.

**Table 2.** Sensitivity of the sex effects to model specification. Women − men differences in the rate of BAG change, in BAG-years per year; positive values indicate faster BAG increase in women. Sex × time is estimated across all participants; sex × amyloid × time is the difference between the sex effect among amyloid-positive and among persistently amyloid-negative participants. The primary model adjusts for age, education and ICV, with education × time and ICV × time. The age-only comparator omits education, ICV and their interactions with time; each remaining sensitivity row changes one element of the primary model. n, participants in the whole-sample / interaction analyses. *Field strength is confounded with ADNI phase (ADNI-1 scanned at 1.5 T), so effects of field strength and study phase cannot be separated by this adjustment.

| Specification | n | Sex × time, all participants [95% CI] | p | Sex × amyloid × time [95% CI] | p |
| --- | --- | --- | --- | --- | --- |
| Primary model | 1,337 / 1,280 | 0.125 [0.031, 0.219] | 0.009 | 0.261 [0.084, 0.438] | 0.004 |
| Age as the only covariate (comparator) | 1,337 / 1,280 | 0.141 [0.054, 0.227] | 0.001 | 0.260 [0.083, 0.436] | 0.004 |
| + Age × time | 1,337 / 1,280 | 0.044 [–0.043, 0.131] | 0.326 | 0.265 [0.110, 0.419] | <0.001 |
| Without ICV | 1,337 / 1,280 | 0.140 [0.052, 0.228] | 0.002 | 0.261 [0.084, 0.437] | 0.004 |
| + Baseline diagnosis × time | 1,337 / 1,280 | 0.187 [0.096, 0.279] | <0.001 | 0.266 [0.094, 0.437] | 0.002 |
| + Baseline ADAS-Cog-13 × time | 1,337 / 1,280 | 0.184 [0.091, 0.277] | <0.001 | 0.241 [0.067, 0.415] | 0.007 |
| + APOE $\epsilon 4$ allele dose × time | 1,337 / 1,280 | 0.134 [0.043, 0.224] | 0.004 | 0.284 [0.111, 0.458] | 0.001 |
| + MRI field strength × time* | 1,337 / 1,280 | 0.127 [0.033, 0.220] | 0.008 | 0.257 [0.081, 0.433] | 0.004 |

Additional model-specification analyses, analyses of clinical progression, detailed centering procedures, and technical model-development analyses are reported in the Supplementary Methods.

## Results

### Brain Age Model Accuracy

On held-out UK Biobank data, the ensemble base model achieved a mean absolute error (MAE) of 2.08 years, compared with 2.15 years after adaptation using MCSA and OASIS-3. Across the MCSA and OASIS-3 validation sets, the adapted model achieved an MAE of 2.65 ± 0.06 years. When applied without further training or fine-tuning to independent cognitively unimpaired, amyloid-negative ADNI participants at baseline, SPARROW achieved an MAE of 2.78 years. Detailed performance across datasets and training stages is shown in Figure 1.

**Figure 1.**
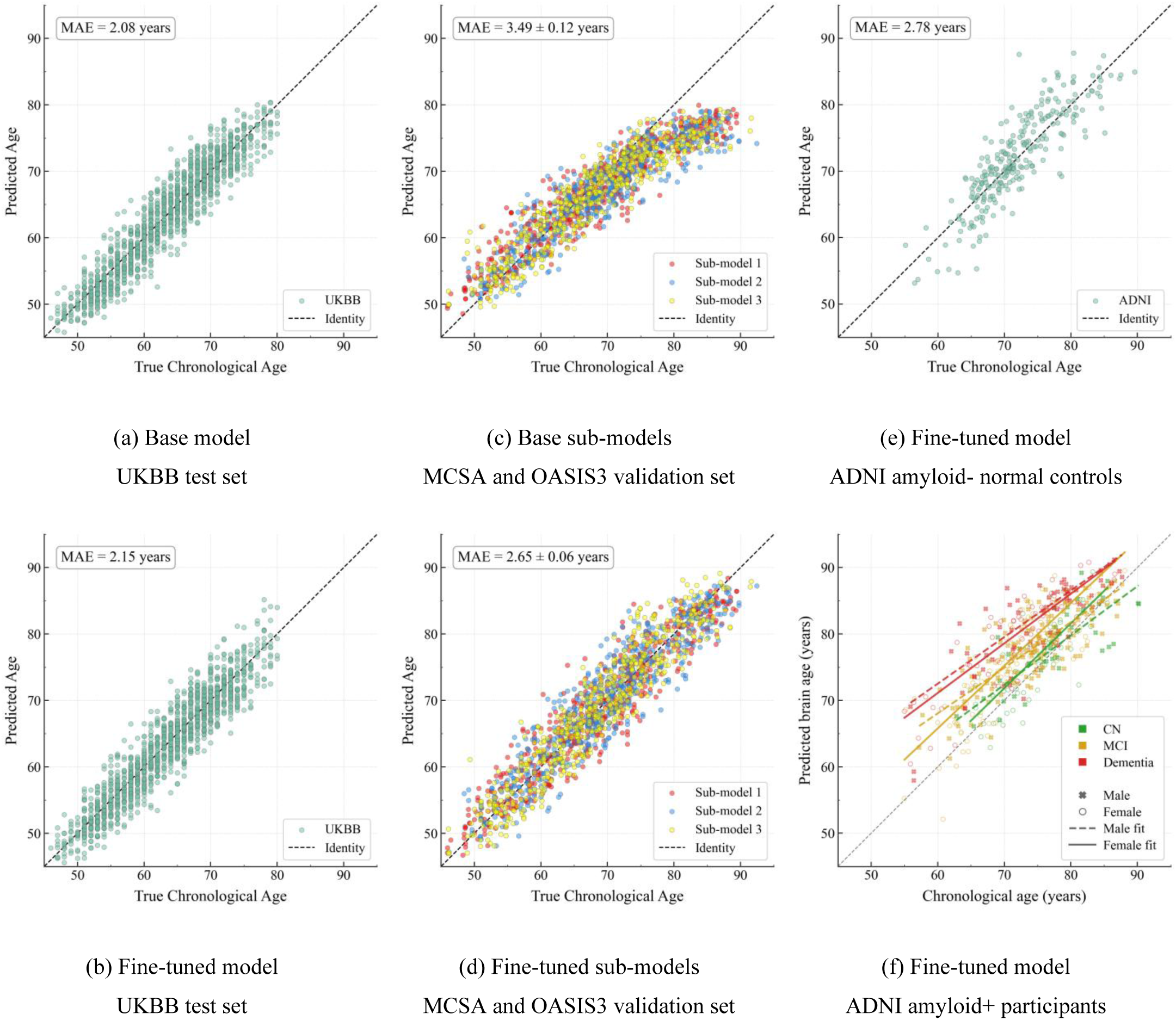
Predicted vs. chronological age for brain age models across datasets and training stages. Each panel shows predicted age on the y-axis and true chronological age on the x-axis, with the dashed line indicating perfect prediction. MAE is reported in each panel. (a) Base model on held-out UKBB neurologically healthy participants. (b) Fine-tuned model on the same held-out UKBB data. (c) Base sub-models (before fine-tuning) on amyloid-negative cognitively healthy participants from MCSA and OASIS3, with each sub-model evaluated on its own validation fold. (d) Fine-tuned sub-models on the same MCSA and OASIS3 validation folds as (c), with each sub-model evaluated on the fold held out from its own adaptation; MAE is the mean ± SD across the three sub-models. (e) Fine-tuned model on amyloid-negative cognitively normal ADNI participants at baseline. (f) Fine-tuned model on amyloid-positive ADNI participants at baseline, colored by clinical status (CN, MCI, dementia), with sex-specific scatter markers and regression lines. Summary provided beside each sub-figure.

### The Analysis Sample

The analysis sample included 1,337 ADNI participants contributing 4,399 MRI visits, with a mean of 3.3 visits per participant over 3.1 years of follow-up. At baseline, 719 participants (53.8%) were amyloid-positive and 618 were amyloid-negative.

Compared with amyloid-negative participants, amyloid-positive participants were older (73.9 ± 7.0 vs 71.5 ± 7.3 years), slightly less educated (15.9 ± 2.8 vs 16.3 ± 2.6 years, p = 0.004), more frequently APOE ε4 carriers (64.3% vs 19.9%) or homozygotes (15.9% vs 1.1%), and more cognitively impaired (ADAS-Cog-13, 19.5 ± 10.6 vs 10.9 ± 6.4). Mild cognitive impairment or dementia was also more frequent among amyloid-positive participants (77.8% vs 44.0%), and mean BAG was greater (3.7 ± 4.6 vs 1.0 ± 3.9 years; all p < 0.001 unless otherwise specified).

Sex was balanced overall and within amyloid strata: women represented 48.2% of the full sample, 50.0% of amyloid-negative participants, and 46.6% of amyloid-positive participants (p = 0.235). Amyloid positivity did not differ significantly between women and men among cognitively normal participants (33.0% vs 29.8%, p = 0.50) or those with mild cognitive impairment (57.6% vs 61.6%, p = 0.32) and was numerically higher in women with dementia (92.9% vs 83.9%, p = 0.058). APOE ε4 carriage was also similar between women and men within each amyloid stratum (amyloid-negative, 20.1% vs 19.7%; amyloid-positive, 64.8% vs 63.8%).

Men were older than women in both amyloid strata (amyloid-negative, 72.4 vs 70.5 years; amyloid-positive, 75.1 vs 72.4 years) and had approximately one additional year of education (16.7 vs 16.0 and 16.3 vs 15.4 years, respectively). Age, education, and ICV were therefore included as covariates in all longitudinal models. Baseline BAG did not differ appreciably by sex in either stratum (amyloid-negative, 0.8 vs 1.2 years; amyloid-positive, 3.9 vs 3.4 years; women vs men).

### Primary analyses

#### Sex differences in BAG trajectories depend on amyloid status

Women and men did not differ in BAG at baseline, either in the whole sample (0.01 years [95% CI −0.54, 0.56], p = 0.98) or within amyloid strata (both p ≥ 0.31).

The primary sex × amyloid status × time interaction was significant (β = −0.261 BAG-years/year [−0.438, −0.084], p = 0.004; n = 1,280), indicating that longitudinal sex differences in BAG varied according to amyloid status.

Among amyloid-positive participants (n = 719), BAG increased by 0.455 years/year in women and 0.176 years/year in men, corresponding to a sex difference of 0.279 BAG-years/year [0.154, 0.404] (p < 0.001). Among amyloid-negative participants who remained amyloid-negative during follow-up (n = 561), BAG trajectories were approximately flat in both women and men (−0.056 and −0.074 years/year, respectively), with a sex difference of 0.018 years/year [−0.118, 0.154] (p = 0.794; Figure 2A–B).

**Figure 2.**
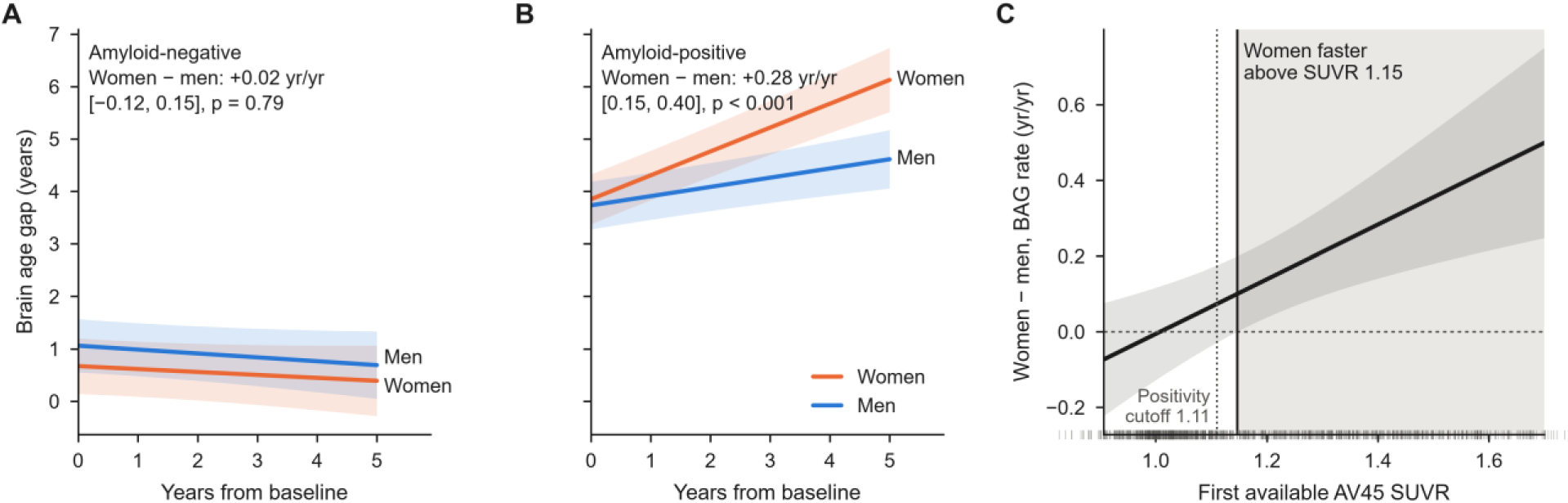
Sex differences in BAG trajectories by amyloid status. (A, B) Model-predicted BAG over follow-up in women and men among persistently amyloid-negative (A) and amyloid-positive (B) participants, from the sex × amyloid × time model evaluated at the sample’s average covariate profile, with 95% confidence bands; the inset gives the women − men difference in BAG rate. (C) Johnson–Neyman plot: the women−men difference in the rate of BAG change as a function of amyloid burden at the first available AV45 assessment (SUVR), with 95% confidence band. The difference is statistically distinguishable from zero in the shaded region, above SUVR 1.146; the dotted line marks the conventional positivity threshold of 1.11. Ticks along the x-axis show individual first available AV45 SUVR values.

The pattern was also observed when amyloid burden was modeled continuously. Each SD increase in florbetapir (AV45) SUVR at the first available assessment was associated with an additional 0.166 BAG-years/year [0.066, 0.265] in the sex difference in BAG change (p = 0.001; n = 910). CSF Aβ42 showed the corresponding association in the expected inverse direction (p = 0.002; n = 924). When amyloid and CSF p-tau181 were included simultaneously as moderators, the amyloid interaction remained significant (p = 0.031), whereas the p-tau interaction was not (p = 0.149).

In an exploratory Johnson–Neyman analysis using first available AV45 measurements, the sex difference in BAG change became statistically distinguishable from zero at an AV45 SUVR of 1.146, slightly above the conventional positivity threshold of 1.11. Detailed results across the continuous amyloid distribution are shown in Figure 2C.

#### APOE ε4 shows an allele dose-dependent association with BAG

APOE ε4 was associated with both baseline BAG and longitudinal BAG change in an allele dose-dependent manner. Relative to participants without an ε4 allele, participants with one ε4 allele had a baseline BAG 1.816 years higher [1.312, 2.319] and a BAG rate 0.258 years/year higher [0.168, 0.347], whereas participants with two ε4 alleles had a baseline BAG 3.325 years higher [2.487, 4.163] and a BAG rate 0.591 years/year higher [0.428, 0.753] (all p < 0.001). The additional effect of the second ε4 allele was significant for both baseline BAG and longitudinal change (p = 0.001 and p < 0.001, respectively; Figure 3).

**Figure 3.**
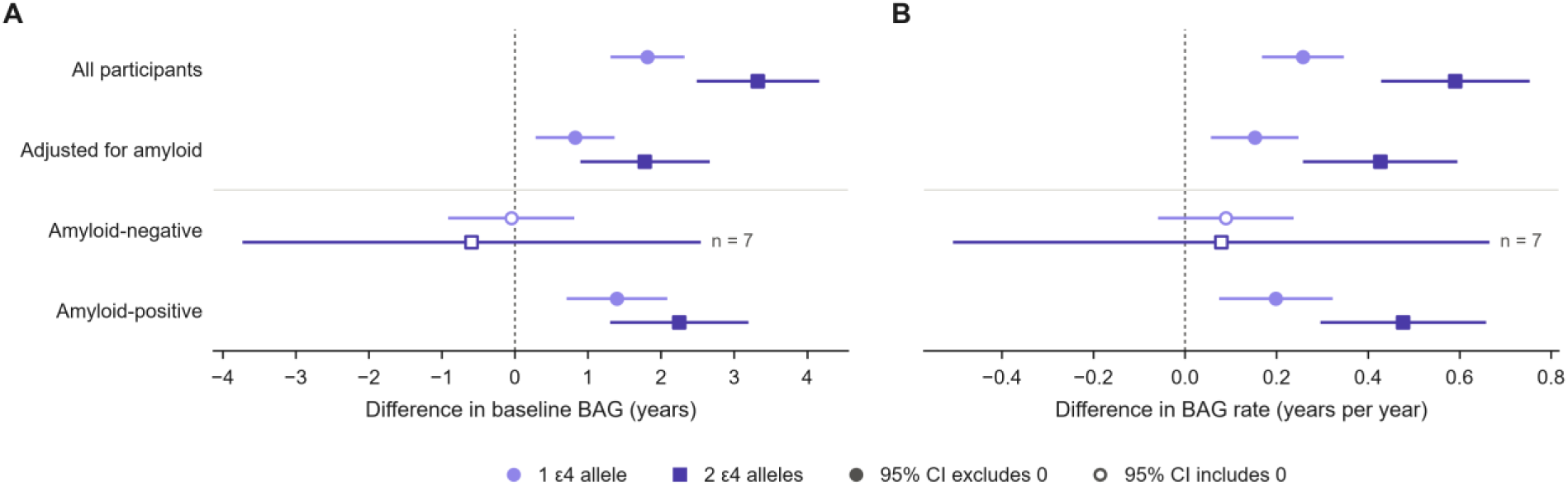
APOE ε4 allele dose and BAG. Difference in baseline BAG (A) and in BAG rate (B) for one (circles) and two (squares) ε4 alleles relative to none: in all participants, after adjustment for baseline amyloid status, and within participants amyloid-negative and amyloid-positive at baseline (contrasts from a single ε4 × amyloid × time model). Hollow markers indicate 95% CIs that include zero. Only seven amyloid-negative participants carried two ε4 alleles.

Adjustment for baseline amyloid status substantially attenuated the APOE ε4 associations. Within amyloid-positive participants, carriers of one ε4 allele had a 1.397-year higher BAG and 0.199 BAG-years/year faster increase, whereas ε4 homozygotes had differences of 2.250 years and 0.477 BAG-years/year, respectively (all p ≤ 0.002). Among amyloid-negative participants, neither one ε4 allele nor ε4 homozygosity was significantly associated with BAG level or rate of change (Figure 3).

However, formal APOE ε4 × amyloid interactions provided more limited evidence of heterogeneity by amyloid status. The interaction for baseline BAG was significant among one-allele carriers (p = 0.010), whereas the interaction for longitudinal BAG change was not (p = 0.268). Interactions among ε4 homozygotes were also not significant, although only seven amyloid-negative participants carried two ε4 alleles. Thus, the allele dose-response association with BAG was clear, whereas evidence that the longitudinal APOE ε4 association differed by amyloid status was limited.

### Secondary analyses

#### Anatomical correlates of BAG

Higher BAG was associated with greater structural brain degeneration across the six volumetric measures examined: hippocampus, entorhinal cortex, middle temporal gyrus, fusiform gyrus, whole brain, and ventricles.

Between-person associations were strongest for hippocampal volume (β = 0.379 per SD [0.345, 0.413]) and weakest for ventricular enlargement (β = 0.092 [0.057, 0.127]); all p < 0.001.

Within-person increases in BAG were also associated with structural change across all six volumetric measures (β = 0.058–0.123; all p < 0.001), indicating that increasing BAG tracked longitudinal tissue loss.

Among persistently amyloid-negative participants, within-person BAG associations remained significant for the hippocampus, fusiform gyrus, whole brain, and ventricles (all p < 0.001). Between-person associations were stronger among amyloid-positive participants for five of the six structures (BAG × amyloid, FDR-adjusted p < 0.016; Figure 4A). The only significant within-person BAG × amyloid interaction after false-discovery-rate correction was for the entorhinal cortex (β = 0.132, FDR-adjusted p = 0.015), where the within-person association was present among amyloid-positive participants (β = 0.083, p = 0.004) but not among amyloid-negative participants (β = −0.030, p = 0.456).

**Figure 4.**
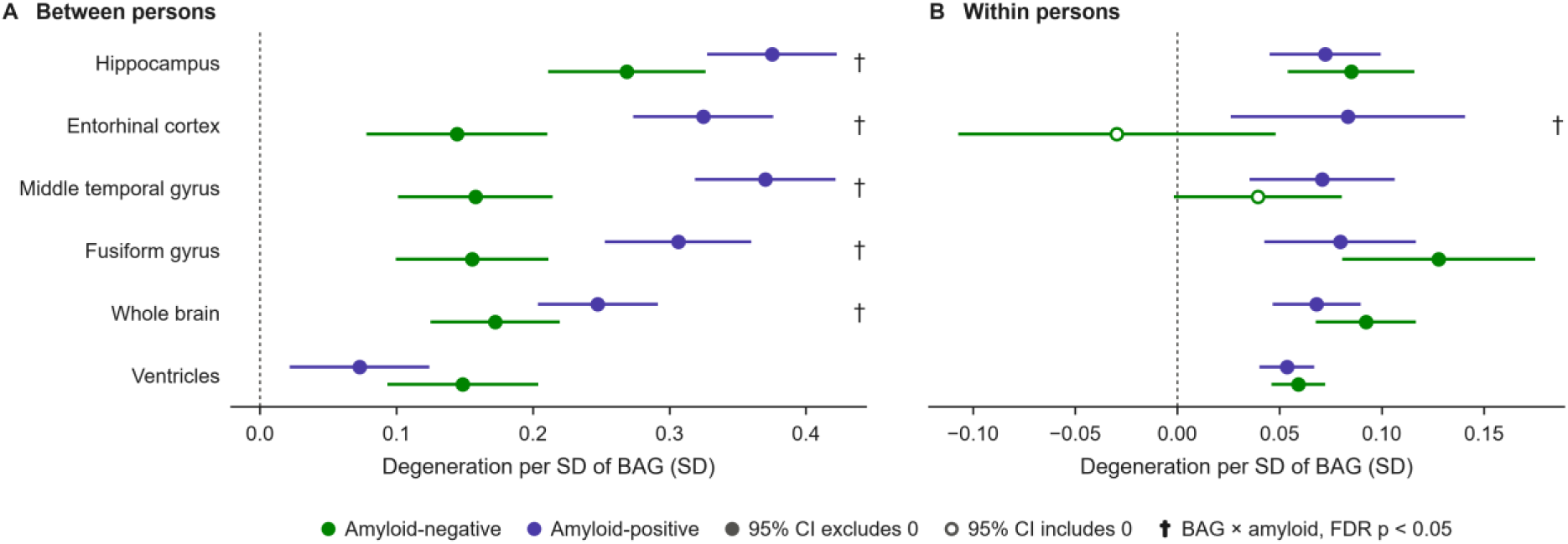
Anatomical correlates of BAG. Standardized associations between BAG and six volumetric measures, decomposed into between-person (a participant’s mean BAG) and within-person (each visit’s deviation from that mean) components, in persistently amyloid-negative and amyloid-positive participants, adjusted for age, sex, education, intracranial volume and time. All outcomes are oriented so that positive means more degeneration. † BAG × amyloid interaction, FDR-adjusted p < 0.05. Hollow markers indicate 95% CIs that include zero.

Anatomical associations were also broadly similar between women and men. Among 12 BAG × sex tests, only the between-person ventricular association survived false-discovery-rate correction (FDR-adjusted p = 0.030).

#### Associations with amyloid and p-tau

Among participants with both AV45 PET amyloid and CSF p-tau181 measurements available (n = 683), both biomarkers were associated with BAG level and longitudinal change. Per SD increase in amyloid burden, BAG was 1.215 years higher at baseline [0.838, 1.592] and increased by an additional 0.112 years/year [0.047, 0.178]. Per SD increase in p-tau, BAG was 0.747 years higher [0.369, 1.125] and increased by an additional 0.188 years/year [0.123, 0.254] (all p < 0.001).

The amyloid and p-tau coefficients did not differ significantly for either baseline BAG (p = 0.169) or longitudinal BAG change (p = 0.195). Because amyloid and p-tau were strongly correlated and biomarker-discordant participants were uncommon, these analyses were not designed to distinguish their relative contributions.

#### BAG–cognition associations differ by amyloid status

Between-person associations between BAG and cognition were present in both amyloid groups across all five cognitive measures. Within-person associations differed more clearly by amyloid status.

Among amyloid-positive participants, within-person increases in BAG were associated with worsening performance on all five cognitive measures (β = 0.075–0.181; all p ≤ 0.020). Among persistently amyloid-negative participants, BAG change was not associated with ADAS-Cog-13, MMSE, RAVLT immediate recall, or MoCA (|β| ≤ 0.019). Equivalence testing against prespecified bounds of ±0.10 SD supported effects equivalent to zero for all four measures (Figure 5).

**Figure 5.**
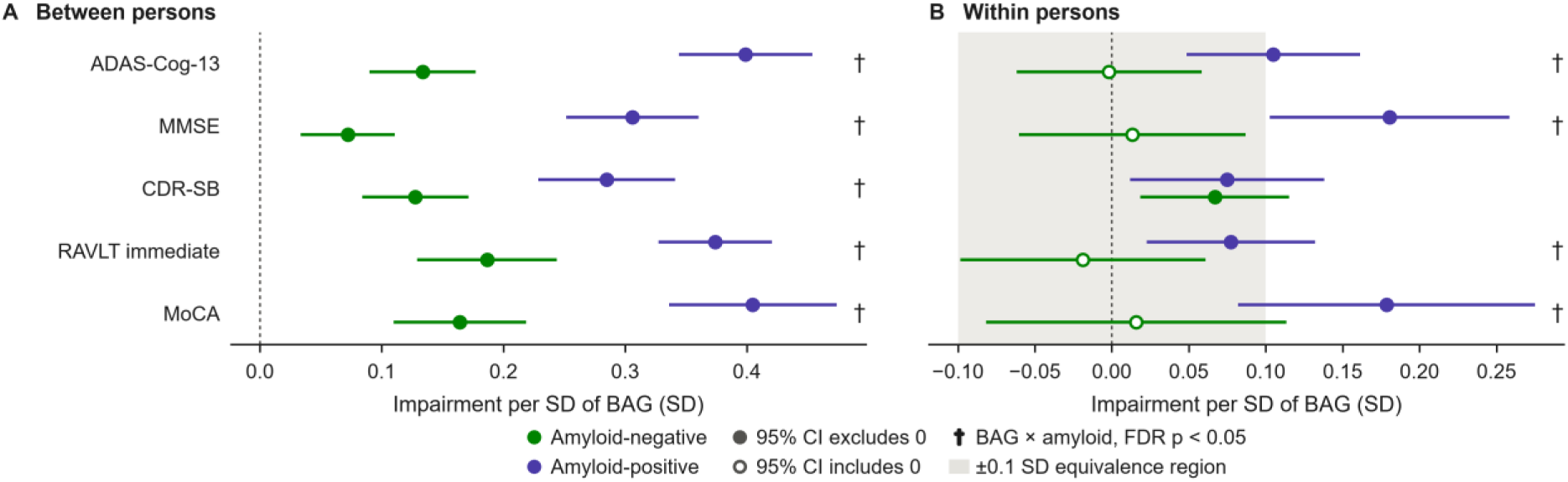
BAG–cognition associations by amyloid status. Between-person (A) and within-person (B) associations between BAG and five cognitive measures, oriented so that positive values indicate more impairment, in persistently amyloid-negative and amyloid-positive participants, adjusted for age, sex, education, intracranial volume and time. Estimates are expressed as cognitive-score SD per SD of BAG, with 95% confidence intervals. The shaded region in B marks the ±0.10 SD equivalence bounds. Equivalence was assessed using two one-sided tests at α = 0.05, corresponding to a 90% confidence interval lying within these bounds. † BAG × amyloid interaction, FDR-adjusted p < 0.05. Hollow markers indicate 95% CIs that include zero.

CDR Sum of Boxes was the exception, showing within-person associations with BAG in both amyloid-positive (β = 0.075) and amyloid-negative participants (β = 0.067).

Formal BAG × amyloid interactions were significant for four of the five within-person cognitive associations (β = 0.144–0.237; all FDR-adjusted p ≤ 0.004), with CDR Sum of Boxes again the exception (β = 0.042, p = 0.361). None of the ten BAG × sex interactions survived correction for multiple comparisons (smallest FDR-adjusted p = 0.066).

#### No significant association between baseline BAG and subsequent amyloid positivity

Among participants who were amyloid-negative at baseline and had at least two amyloid assessments (n = 442; 57 conversions), baseline BAG was not significantly associated with subsequent amyloid positivity (HR 0.958 per SD [0.744, 1.233], p = 0.739). The estimate was similar in the unadjusted model (HR 0.980, p = 0.876).

In the same model, APOE ε4 carrier status predicted subsequent amyloid positivity (HR 2.342 [1.333, 4.114], p = 0.003). Neither sex (HR 0.636 [0.334, 1.211], p = 0.168) nor the BAG × sex interaction (p = 0.426) was significant. Because the number of amyloid conversions was limited, the BAG result should be interpreted as absence of a detected association rather than evidence of equivalence.

#### BAG predicts clinical progression

Higher baseline BAG was associated with subsequent clinical progression. Among participants with mild cognitive impairment at baseline, each SD higher BAG was associated with greater risk of progression to dementia (HR 1.667 [1.460, 1.904], p < 0.001; n = 594, 216 events; Supplementary Figure S2). Among cognitively normal participants, BAG was also associated with progression to cognitive impairment (HR 1.397 [1.136, 1.719], p = 0.002; n = 487, 84 events).

The BAG × sex interaction was not significant for either transition (p = 0.060 and p = 0.142).

Among persistently amyloid-negative participants with mild cognitive impairment, 28 of 221 progressed to dementia. Baseline BAG remained associated with progression in this group (HR 1.659 [1.132, 2.433], p = 0.010), similar to the estimate among amyloid-positive participants (HR 1.672 [1.444, 1.936]). BAG × amyloid interactions were not significant for either clinical transition (p = 0.712 and p = 0.902), although the small number of events among amyloid-negative participants limited precision.

Sex itself was not significantly associated with clinical progression among participants with CN or MCI at the first retained MRI scan (HR 1.130 [0.848, 1.505], p = 0.404), among amyloid-positive participants (HR 1.031 [0.737, 1.442], p = 0.858), or among amyloid-positive participants with mild cognitive impairment at baseline (HR 0.924 [0.634, 1.348], p = 0.682). These hazard ratios compare men with women.

### Sensitivity analyses

The sex × amyloid status × time interaction was stable across model specifications (Table 2). The women − men difference in BAG rate among amyloid-positive relative to amyloid-negative participants was 0.24–0.28 BAG-years/year (all p ≤ 0.007) when baseline diagnosis × time, baseline ADAS-Cog-13 × time, APOE ε4 allele dose × time, MRI field strength × time, or age × time was added, or when ICV was removed. Adding age × time, which allows baseline age to be associated with longitudinal BAG change, yielded an interaction similar in magnitude (0.265 [0.110, 0.419], p < 0.001) but reduced the sex difference in BAG rate across the whole sample from 0.125 [0.031, 0.219] (p = 0.009) to 0.044 [−0.043, 0.131] (p = 0.33).

Restricting the continuous AV45 analysis to measurements recorded at the study baseline exam yielded 728 participants and 1,989 MRI visits. The women − men sex × AV45 × time interaction remained positive but its 95% confidence interval included zero (β = 0.121 BAG-years/year per SD higher AV45 [95% CI −0.013, 0.254], p = 0.077). No Johnson–Neyman significance boundary was identified within the observed SUVR range. This restriction excluded 182 participants, including 180 originally enrolled in ADNI-1.

## Discussion

### Principal findings

Our findings suggest that BAG should be interpreted in the context of amyloid status. BAG tracked structural neurodegeneration in both amyloid-positive and amyloid-negative participants, but its relationships with cognition and established AD risk factors were more evident among amyloid-positive participants.

Within individuals, increasing BAG was accompanied by loss of hippocampal, fusiform, and whole-brain tissue irrespective of amyloid status, and the anatomical correlates of BAG were broadly similar in women and men. The main exception was the entorhinal cortex, an early vulnerable region in AD, where the within-person association with BAG was present among amyloid-positive but not amyloid-negative participants.

Among participants who remained amyloid-negative, within-person BAG change was unrelated to change on four of five cognitive measures, with all four effects equivalent to zero within ±0.1 SD. Baseline BAG also did not predict subsequent amyloid positivity, whereas APOE ε4 did in the same model. However, higher baseline BAG predicted clinical progression, including progression from mild cognitive impairment to dementia. Thus, BAG remained clinically relevant in amyloid-negative participants but was not specifically associated with subsequent amyloid emergence.

Sex differences were primarily longitudinal and depended on amyloid status. Women and men did not differ in BAG at baseline, but BAG increased faster in women among amyloid-positive participants, whereas trajectories were similar in persistently amyloid-negative participants. The sex × amyloid × time interaction was significant. When amyloid burden was analyzed continuously using first available AV45 measurements, the sex difference became statistically distinguishable from zero at an AV45 SUVR of 1.146, slightly above the conventional positivity threshold of 1.11. This threshold was exploratory, was not identified in the baseline-only sensitivity analysis, and requires replication.

APOE ε4 was associated with BAG in an allele dose-dependent manner at both baseline and longitudinally, with the second allele adding significantly to the first. Previous brain-age studies have generally dichotomized APOE ε4 or compared genotypes without explicitly testing an ordered allele-dose effect^30–32^. Adjustment for amyloid status substantially attenuated the APOE ε4 associations, and the associations remained evident among amyloid-positive participants. However, formal APOE ε4 × amyloid interactions provided more limited evidence that the longitudinal APOE ε4 association itself differed by amyloid status, particularly because only seven amyloid-negative participants were ε4 homozygotes. We therefore interpret the allele-dose association as robust, while the extent to which it depends on amyloid status remains less certain.

These findings may help reconcile apparently conflicting reports of sex differences in brain aging. Taylor et al.^7^ reported faster BAG increase in women in ADNI, particularly in comparisons involving progressive mild cognitive impairment, whereas Cumplido-Mayoral et al.^3^ reported higher BAG in cognitively unimpaired A+T+ women, although the formal sex-by-biomarker interaction was not significant. In contrast, Millar et al.^8^ reported higher BAG in men in autosomal-dominant AD, but the difference was also present in non-carriers and did not change with proximity to symptom onset. Ravndal et al.^11^ similarly reported faster brain aging in cognitively healthy men. Taken together, these studies and our findings suggest that the direction and magnitude of sex differences in brain aging may vary with disease stage and biomarker context.

Our findings differ from reports from the Baltimore Longitudinal Study of Aging. Armstrong et al.^9^ found stronger amyloid-related longitudinal atrophy in cognitively normal men than women, and Joynes et al.^10^ reported steeper post-amyloid-onset declines in brain volume and cognition among men. Those cohorts were smaller and predominantly preclinical, whereas most amyloid-positive participants in our sample had mild cognitive impairment or dementia. Differences in disease stage, cohort composition, and analytic approach may therefore contribute to the differing results. Other studies have also located sex differences downstream of amyloid, either through interactions with amyloid burden^33,34^ or sex differences in tau accumulation measured by PET after accounting for amyloid^35^.

### What We Did Not Find

Several null findings further constrain interpretation. We found no baseline sex difference in BAG, indicating that the principal sex effect was longitudinal rather than cross-sectional. Education was not associated with longitudinal BAG change (education × time, p = 0.43), despite its established relationship with cognitive reserve ^36,37^. Sex was also not significantly associated with clinical progression, despite faster BAG increase in amyloid-positive women. Thus, the observed sex difference in structural brain aging did not translate into detectable earlier clinical progression over the available follow-up.

Baseline BAG did not predict subsequent amyloid positivity, although this analysis was not sufficiently precise to establish equivalence. We also found no evidence that the BAG–cognition relationship differed by sex after correction for multiple comparisons. Finally, both amyloid and p-tau were associated with BAG level and longitudinal change, but their associations were not statistically distinguishable. Because these biomarkers were strongly correlated in ADNI and biomarker-discordant participants were uncommon, the present analyses cannot separate their relative contributions.

### Limitations

This work is exploratory and has no external replication. Effects were tested across many domains without correcting p-values between them. The sex × amyloid × time interaction is the only result we would call confirmatory in intent; everything else, including the Johnson–Neyman threshold, is hypothesis-generating. The sample is restricted to ADNI, whose participants are predominantly White, highly educated and clinic-recruited, so both the threshold and the effect size may differ elsewhere.

Several analyses are underpowered, and where we report that an interaction was null the honest reading is a failure to detect rather than a demonstration of equality. The clinical prediction in amyloid-negative participants rests on 28 events, and only 7 amyloid-negative participants carry two ε4 alleles.

The AV45 measurement was recorded after the study baseline exam for 20% of participants, so this analysis does not establish that amyloid burden measured at study entry predicts subsequent sex differences in BAG trajectories. The interaction was not statistically significant and no Johnson–Neyman boundary was identified when AV45 was restricted to study baseline. That restriction also removed all 180 participants originally enrolled in ADNI-1 from the AV45 sample and reduced longitudinal coverage, so the sensitivity result cannot be attributed to measurement timing alone.

Two further caveats. MRI field strength was correlated with other covariates and confounded with ADNI phase, so it was omitted from the primary model. A sensitivity analysis added field strength × time (Table 2). This adjustment cannot separate field-strength effects from differences associated with ADNI phase, and residual scanner-related effects cannot be ruled out. One sensitivity analysis adjusts for baseline ADAS-Cog-13, which is partly downstream of the tissue loss BAG measures. We report it because the first objection to a sex difference in a clinical cohort is that women were more impaired to begin with, and adjusting for severity addresses that objection directly; but because the adjustment also removes part of the effect being estimated, that estimate should be read as a conservative one.

### Future Work

Replication should prioritize cohorts with longitudinal MRI and amyloid measurements spanning preclinical and symptomatic stages. In particular, applying the same sex × amyloid × time model in cohorts that have reported sex effects in the opposite direction would test whether differences in amyloid status or disease stage account for the apparent inconsistency. The exploratory AV45 threshold should also be evaluated in cohorts with prospectively aligned baseline AV45 and MRI assessments and different amyloid distributions.

Longer follow-up will be needed to determine whether faster BAG increase in amyloid-positive women eventually translates into differences in clinical progression. More broadly, our findings suggest that BAG should be interpreted alongside biomarker status rather than as an AD-specific marker in isolation. In biomarker-negative populations, elevated BAG may identify structural brain aging and clinical vulnerability without necessarily indicating emerging amyloid pathology.

## Data Availability

All data produced in the present study are available upon reasonable request to the authors.

