## Supplementary Methods for "Sex and APOE ε4 allele dose relate to longitudinal brain aging by amyloid status"

#### Cohort selection and MRI acquisition

##### UK Biobank

The UK Biobank (UKBB) is a large-scale population resource designed to investigate determinants of human health using imaging, genetic, clinical, and other phenotypic data[1]. We used UKBB imaging release v14940, which included 39,676 T1-weighted MRI scans. Images were acquired on 3T Siemens Skyra scanners using a 3D MPRAGE sequence (TR = 2000 ms, TE = 2.01 ms, TI = 880 ms, flip angle = 8°, 1 $mm^3$ isotropic voxels)[2,3].

To train the base brain-age model on brains without known neurological or structural abnormalities, we excluded participants with infectious or parasitic diseases involving the central nervous system; neoplasms involving the brain, meninges, or cranial nerves; metabolic or nutritional disorders known to alter brain structure; organic mental disorders; diseases of the nervous system; cerebrovascular disease; congenital malformations affecting the central nervous system; head injuries; poisonings and related sequelae; external-cause codes indicating structural brain injury; or abnormal imaging or laboratory findings suggestive of structural brain change. After these exclusions and quality control, 26,512 participants were included in base-model training.

##### Mayo Clinic Study of Aging

The Mayo Clinic Study of Aging (MCSA) is a longitudinal, population-based study of aging and cognitive decline among community-dwelling adults in Olmsted County, Minnesota[4]. We included 1,297 T1-weighted MRI scans from 1,123 cognitively unimpaired, amyloid-negative participants.

Imaging was performed on 3T GE scanners using a 3D T1-weighted sequence (TR = 6.6–7.4 ms, TE = 2.8–3.0 ms, TI = 900 ms, flip angle = 8°, voxel size = 1 × 1 × 1.2 mm) and 3T Siemens Prisma scanners using a 3D MPRAGE sequence (TR = 2300 ms, TE = 3.1 ms, TI = 945 ms, flip angle = 9°, voxel size = 0.8 × 0.8 × 0.8 mm)[4]. Amyloid status was determined using PET data available within the cohort.

MCSA was used together with OASIS-3 to adapt the pretrained brain-age model to older ages and imaging environments that differed from those represented in UKBB.

#### Open Access Series of Imaging Studies

OASIS-3 is an open-access longitudinal neuroimaging resource containing data acquired across adulthood and older age over approximately three decades[5,6]. We included 2,035 T1-weighted MRI scans from 654 cognitively normal, amyloid-negative participants.

MRI was performed on 3T Siemens TIM Trio scanners using a 3D MPRAGE sequence (TR = 2400 ms, TE = 3.08 ms, TI = 1000 ms, flip angle = 8°, 1 $mm^3$ isotropic voxels). Amyloid status was determined from PET data available within OASIS-3. OASIS-3 and MCSA together constituted the intermediate model-adaptation sample.

#### Alzheimer's Disease Neuroimaging Initiative

The Alzheimer's Disease Neuroimaging Initiative (ADNI) is a multisite longitudinal public–private partnership established to identify imaging, clinical, and biological markers of Alzheimer's disease progression. We included 3D T1-weighted MRI from participants enrolled across ADNI-1, ADNI-Go, ADNI-2, ADNI-3, and ADNI-4.

Images were acquired across multiple sites using 1.5T scanners in ADNI-1 and predominantly 3T scanners in ADNI-Go, ADNI-2, ADNI-3, and ADNI-4. Scanners from GE, Siemens, and Philips were used with standardized 3D MPRAGE or vendor-equivalent inversion-recovery spoiled gradient-echo protocols. Representative acquisition parameters were TR ≈ 2300 ms, TE ≈ 3 ms, TI ≈ 900–1000 ms, flip angle ≈ 8–9°, and near-isotropic voxel dimensions of approximately 1.0–1.1 × 1.0–1.1 × 1.2 mm for ADNI-1/Go/2 and 1.0 × 1.0 × 1.0 mm for ADNI-3/4.

No ADNI images were used during training or adaptation of the brain-age model. Amyloid-negative, cognitively unimpaired ADNI participants were first used as an independent evaluation sample, after which the fixed model was applied to the complete ADNI analysis cohort.

#### Amyloid classification

Amyloid status in the MCSA and OASIS-3 model-adaptation samples was determined from the PET components of the respective cohorts.

### SPARROW brain-age architecture

We refer to the brain-age model as SPARROW: Simple Pseudo-fully-convolutional Architecture with Residuals, Regularized, Optimized and Weight-consolidated. SPARROW was derived from the SFCN-reg backbone used in our previous work[7], with modifications intended to improve training stability, regularization, and adaptation across datasets.

#### Residual architecture

The network contains six residual blocks with 32, 64, 128, 256, 256, and 128 filters, respectively. Each block consists of:

Convolution → Group Normalization → ReLU → Convolution → Group Normalization → ReLU → Pooling.

The residual connection links the input of the first convolution directly to the output of the second Group Normalization layer. An identity connection is used when the number of input and output channels is unchanged; otherwise, a $1 \times 1 \times 1$ convolution is used to match the number of feature channels.

Residual connections were introduced after observing instability in the loss curves of the preceding architecture. Reformulating the learned mapping as a residual function added to an identity shortcut can facilitate optimization of deep neural networks[8].

#### Stochastic-depth and dropout regularization

Stochastic depth[9], implemented as DropPath, was applied to residual branches during training. The DropPath probability increased linearly across the network, from 0 in the first block to 0.10 in the final block. This approach preserves early feature extraction while increasing regularization in deeper layers[10].

The final residual block uses three-dimensional average pooling rather than max pooling to reduce each feature map to a single value. This yields a 128-dimensional feature representation, which is passed through a dropout layer with probability 0.20 before the final prediction layer.

#### Group Normalization

The addition of a second convolution to each residual block increased memory requirements and reduced the feasible training batch size. Because Batch Normalization may become unstable or less effective at small batch sizes, it was replaced with Group Normalization, whose normalization statistics do not depend directly on batch size[11].

#### Exponential moving average

To reduce variability caused by stochastic gradient updates and obtain a smoother estimate of the final model parameters, we maintained an exponential moving average (EMA) of model weights during training (as defined in Eq. 1). EMA acts directly in weight space and complements DropPath and dropout regularization[12]. The EMA decay coefficient was set to 0.998.

Eq. 1 $$\theta_{EMA} \leftarrow \beta\theta_{EMA} + (1 - \beta)\theta$$

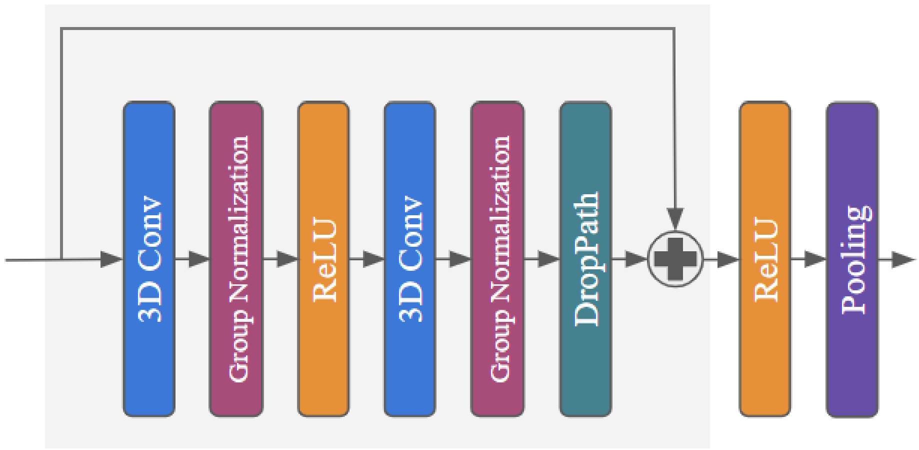


**Supplementary Figure S1.** Architecture of the SPARROW residual block. The model contains six residual blocks. Max pooling with stride 2 is used in the first five blocks; the final block uses average pooling to produce a 128-dimensional feature representation before dropout and age prediction.

##### Ensemble prediction

To reduce model-specific variation arising from random initialization, three independently initialized sub-models (ensemble members) were trained, and final brain-age estimates were obtained by averaging their predictions at inference.

The resulting architecture contained approximately 7.18 million trainable parameters.

#### Continual-learning adaptation to older adults

The UKBB training sample contained relatively few individuals at the oldest ages represented in the ADNI cohort. Directly applying the pretrained model to older participants and different scanner environments therefore risked age-related extrapolation error and domain shift. Conversely, unconstrained fine-tuning on a new dataset can alter previously learned representations and degrade performance on the original training distribution.

We therefore adapted the pretrained model using cognitively unimpaired, amyloid-negative participants from MCSA and OASIS-3. To limit forgetting of the original training domain, UKBB data were retained in the adaptation training sample, and changes to encoder parameters were constrained with Elastic Weight Consolidation.

##### Adaptation data and validation split

MCSA and OASIS-3 participants were assigned to five folds at the participant level, so that all scans from a given participant belonged to the same fold. Each of the three sub-models (see "Ensemble prediction" above) was adapted from its own base model with a different MCSA/OASIS-3 fold held out for validation; the three validation folds did not overlap, and each sub-model was trained on the remaining four folds. Across the three sub-models, the adaptation training sets contained 2,668–2,730 MCSA/OASIS-3 scans and the validation sets contained 601–663 scans (Supplementary Table S1). Pooled across sub-models, the validation folds comprised 756 MCSA scans from 649 participants and 1,115 OASIS-3 scans from 378 participants.

**Supplementary Table S1.** MCSA and OASIS-3 scans used to adapt each sub-model. Values are numbers of scans, with numbers of participants in parentheses. Folds were defined at the participant level, so no participant contributed scans to both training and validation within a sub-model. Age refers to age at the time of the scan.

| | **Sub-model 1** | **Sub-model 2** | **Sub-model 3** |
|---|---|---|---|
| Held-out validation fold (of 5) | Fold 5 | Fold 1 | Fold 2 |
| **Training (remaining four folds)** | | | |
| MCSA | 1,032 (894) | 1,047 (911) | 1,056 (915) |
| OASIS-3 | 1,636 (541) | 1,683 (522) | 1,668 (521) |
| Total | 2,668 (1,435) | 2,730 (1,433) | 2,724 (1,436) |
| Age < 45 years | 3 | 6 | 6 |
| Age 45 to < 81 years | 2,267 | 2,311 | 2,282 |
| Age ≥ 81 years | 398 | 413 | 436 |
| **Validation (held-out fold)** | | | |
| MCSA | 265 (229) | 250 (212) | 241 (208) |
| OASIS-3 | 398 (113) | 351 (132) | 366 (133) |
| Total | 663 (342) | 601 (344) | 607 (341) |
| Age < 45 years | 3 | 0 | 0 |
| Age 45 to < 81 years | 542 | 498 | 527 |
| Age ≥ 81 years | 118 | 103 | 80 |

UKBB data used during adaptation are not shown. Each sub-model used 18 UKBB folds for training (n = 23,862) and a different UKBB fold for validation (n = 1,325); one further UKBB fold (n = 1,325) was held out from all sub-models as the UKBB test set.

UKBB participants had been assigned to 20 folds for base-model training. One fold (n = 1,325) was held out from base-model training and adaptation of all three sub-models and served as the UKBB test set. Each sub-model used a different UKBB fold (n = 1,325) for validation, matching the validation fold of its base model, and the remaining 18 folds (n = 23,862) were included in adaptation training.

#### Prediction-head adaptation

The original linear prediction head was replaced with a multilayer perceptron containing one hidden layer of 64 units. This allowed greater flexibility in the mapping from the learned imaging representation to chronological age across the expanded age range.

#### Linear probing followed by fine-tuning

Adaptation followed a two-stage linear-probing and fine-tuning procedure[13]. During the first five epochs, the convolutional encoder was frozen and only the newly initialized prediction head was trained, using a learning rate of $1 \times 10^{-2}$. This allowed the prediction head to adapt to the new age distribution without altering previously learned imaging features.

During the second phase (epochs 6–40), the encoder was unfrozen. The learning rate was $1 \times 10^{-3}$ for the prediction head and $1 \times 10^{-4}$ for the deepest residual block, halved for each successively shallower block. Optimization used AdamW with a weight decay of $1 \times 10^{-4}$, a cosine learning-rate schedule with linear warm-up over the first 10% of steps, a batch size of 1 with gradients accumulated over 4 steps, and a multi-quantile Huber loss. Dropout before the prediction head was set to 0.30. An exponential moving average (EMA) of model weights with a decay coefficient of 0.999 was maintained and was reinitialized at the start of the second phase.

#### Elastic Weight Consolidation

Elastic Weight Consolidation (EWC)[14] was applied during the second phase to reduce catastrophic forgetting. EWC penalizes changes in parameters that were important for prediction in the original UKBB task. Parameter importance was estimated as the empirical Fisher information of the encoder parameters, computed from 2,000 UKBB training scans, and the penalty anchored encoder parameters to their pretrained values. The prediction head was not constrained. The EWC weight $\lambda$ was initialized at 1,000 and multiplied by 0.9 after each epoch, with a lower bound of 200.

#### Sampling and age imbalance

Training scans were drawn with replacement using a weighted random sampler. Dataset weights of 0.15 for UKBB, 1.25 for MCSA, and 1.0 for OASIS-3 down-weighted the UKBB data relative to the older adaptation cohorts. For each sub-model, the number of draws per epoch was the integer part of the sum of its training-set sizes multiplied by the dataset weights. For sub-model 3, this yielded 6,567 draws per epoch; after applying the LDS, uniform-mixture, and dataset weighting described below, the expected contributions were approximately 2,060 UKBB, 2,209 MCSA, and 2,297 OASIS-3 draws.

The adaptation data were also imbalanced across the age distribution, particularly at the oldest ages. Label Distribution Smoothing (LDS)[15] was therefore used to weight scans by the inverse of a smoothed age-label density (100 age bins; Gaussian kernel of size 5 with $\sigma = 1$; maximum weight 100). LDS weights were mixed equally with uniform weights (temperature 0.5) before the dataset weights were applied.

##### Model selection

Validation mean absolute error (MAE) was computed using EMA weights within three age strata: younger than 45 years, 45 to younger than 81 years, and 81 years or older. Each sub-model's validation set comprised its MCSA/OASIS-3 validation fold and its UKBB validation fold. The model-selection criterion was the weighted sum of the stratum-specific MAEs, with weights of 0.05, 0.40, and 0.55, respectively.

Each sub-model was trained for 40 epochs, and the checkpoint with the lowest weighted validation criterion was retained. The three adapted sub-models were combined by averaging their predictions at inference. Because validation folds differed across sub-models, performance in MCSA/OASIS-3 was evaluated for each sub-model on its own validation fold and summarized across sub-models.

#### Brain age gap calculation

After adaptation to MCSA and OASIS-3, SPARROW was applied to ADNI without additional training or fine-tuning. No ADNI observations contributed to model fitting.

At each MRI visit, brain age gap (BAG) was calculated as:

$$BAG = predicted\ brain\ age - chronological\ age.$$

Positive BAG values therefore indicate that the MRI-derived brain age was older than the participant’s chronological age, whereas negative BAG values indicate a younger-appearing brain.

#### Statistical coding and centering

All categorical predictors were entered using centered contrasts, and continuous predictors were centered. Time from baseline was not centered, so that time = 0 corresponded to the baseline visit.

Sex was coded −0.5 for women and +0.5 for men. Under this coding, the sex coefficient represents the male-minus-female difference at baseline, and a negative sex × time coefficient indicates a faster longitudinal increase in BAG among women.

Covariates were centered using each participant's first retained visit and then averaged across participants so that the covariate origin corresponded to the average baseline profile of the analysis sample. When a model was fitted within a subsample, covariates were re-centered within that subsample.

This parameterization makes lower-order sex terms interpretable as sex differences at the average covariate profile rather than at an arbitrary reference category or at zero values of continuous covariates. Reversing the sex coding changes only the sign of the corresponding coefficients and does not alter their magnitude or statistical significance.

As a sensitivity check, sex coefficients were unchanged to three decimal places when visit-level, participant-level, or baseline centering was used. Highest-order interaction terms, including the primary sex × amyloid × time interaction, are unaffected by the choice of centering.

#### Intracranial-volume adjustment

For analyses in which regional brain volume was the outcome, intracranial volume (ICV) was entered as a covariate rather than using the ratio of regional volume to ICV[16].

Ratio normalization assumes that regional volume changes in direct proportion to head size. When this assumption is not satisfied, dividing by ICV can overcorrect regional volume and leave the resulting ratio associated with ICV. Because head size also differs systematically by sex, ratio normalization can consequently introduce or exaggerate apparent sex differences. Covariate adjustment was therefore used throughout the regional-volume analyses.

#### Mixed-effects model specification

Longitudinal BAG trajectories were estimated using linear mixed-effects models with participant-specific random intercepts and random slopes for time. The random intercept allowed each participant to have a distinct baseline BAG, whereas the random slope allowed individual rates of longitudinal BAG change to vary. Fixed effects in the primary model included sex, baseline age, education, ICV, time, sex × time, education × time, and ICV × time. Baseline age entered as a main effect; age × time was added in a sensitivity analysis. MRI field strength was excluded from the primary model because it was correlated with other covariates.

Models were estimated by maximum likelihood rather than restricted maximum likelihood to allow likelihood-based comparison of models differing in their fixed effects. Statistical analyses were performed in Python using *statsmodels* version 0.15.0[17], with optimization using the Powell algorithm (with L-BFGS as a fallback when the Powell fit did not converge).

When BAG was entered as a longitudinal predictor, it was separated into between-person and within-person components[18], allowing associations attributable to stable differences between individuals to be distinguished from associations attributable to within-person changes over time.

BAG was first standardized using the mean and standard deviation of BAG among cognitively unimpaired, amyloid-negative ADNI participants at baseline (n = 256; 1 SD = 3.64 years). For participant $i$ at visit $j$, let $z_{ij}$ denote standardized BAG, $n_i$ the number of visits contributed by participant $i$, and $N$ the number of participants in the fitted sample. The two components were defined as:

$$BAG_{between_i} = \bar{z}_i - \frac{1}{N}\sum_k \bar{z}_k\,, \qquad where\ \bar{z}_i = \frac{1}{n_i}\sum_j z_{ij}$$

$$BAG_{within_{ij}} = z_{ij} - \bar{z}_i$$

The between-person component is therefore the participant's mean standardized BAG, centred on the mean of participant means, and is constant across a participant's visits. The within-person component is the visit-specific deviation from the participant's own mean and averages to zero within each participant. Both components were computed separately for each fitted model, using only the visits with a non-missing outcome in the sample being analysed.

For regional brain volume and cognitive outcomes, both components were entered simultaneously:

$$Y_{ij} = \beta_0 + \beta_A \cdot age_i + \beta_E \cdot education_i + \beta_V \cdot ICV_i + \beta_S \cdot sex_i + \beta_T \cdot t_{ij} + \beta_B \cdot BAG_{between_i} + \beta_W \cdot BAG_{within_{ij}} + u_{0i} + u_{1i} \cdot t_{ij} + \varepsilon_{ij}$$

where $Y_{ij}$ is the standardized outcome, oriented so that higher values indicate greater degeneration or impairment; $t_{ij}$ is time from baseline in years; and $u_{0i}$ and $u_{1i}$ are participant-specific random intercepts and slopes for time. $\beta_B$ estimates the association between a participant's typical BAG level and the outcome, and $\beta_W$ estimates whether visits with higher-than-usual BAG for that participant coincide with higher outcome values. To test whether these associations differed by amyloid status or sex, each component was additionally multiplied by the centred moderator (amyloid status at the first retained visit, or sex), and the corresponding product terms were tested.

#### Interaction testing and multiple-comparison control

When results were presented separately for amyloid-positive and amyloid-negative participants, evidence that an association differed between groups was evaluated using the corresponding interaction term in the complete analysis sample. Group differences were therefore not inferred solely from an association being statistically significant within one subgroup but not within another.

For families of BAG-by-sex and BAG-by-amyloid interaction tests involving multiple regional or biomarker outcomes, false-discovery-rate correction was applied within each predefined family of tests.

Continuous amyloid analyses were additionally evaluated using the Johnson–Neyman procedure[19,20] to identify the range of amyloid burden over which the estimated sex difference in longitudinal BAG change was statistically distinguishable from zero. Thresholds were expressed both in the original amyloid measurement scale and as percentiles of the observed sample distribution.

#### Equivalence testing

Equivalence testing was used when the scientific question concerned evidence for a negligible association rather than failure to reject a conventional null hypothesis. For cognitive analyses, effects within ±0.10 standard deviations were prespecified as sufficiently small to be considered practically equivalent to zero.

Equivalence was assessed using two one-sided tests (TOST). For an estimated coefficient $\hat{\beta}$ with standard error SE and equivalence margin $\Delta = 0.10$, the null hypotheses $H_{01}: \beta \leq -\Delta$ and $H_{02}: \beta \geq \Delta$ were tested against the alternatives $\beta > -\Delta$ and $\beta < \Delta$, respectively, using Wald $z$ statistics:

$$z_1 = \frac{\hat{\beta} + \Delta}{SE}, \qquad z_2 = \frac{\hat{\beta} - \Delta}{SE}$$

with one-sided p-values $p_1 = 1 - \Phi(z_1)$ and $p_2 = \Phi(z_2)$, where $\Phi$ is the standard normal cumulative distribution function. The TOST p-value was $max(p_1, p_2)$, and equivalence was declared when this value was below 0.05. This criterion is equivalent to the 90% confidence interval, $\hat{\beta} \pm 1.645\, SE$, lying entirely within $(-\Delta, \Delta)$.

Because cognitive outcomes and BAG were both standardized, Δ is expressed in standard deviations of the cognitive outcome per standard deviation of BAG. TOST was applied to the between-person and within-person BAG coefficients from the cognition models fitted separately within amyloid-positive and persistently amyloid-negative participants (see "Mixed-effects model specification" above). Equivalence p-values were not adjusted for multiple comparisons.

### Additional and sensitivity analyses

#### Model-specification sensitivity analyses

The robustness of the two sex estimates (the overall sex × time effect in all participants and the sex × amyloid × time interaction in the primary analysis sample) was assessed by making one change at a time to the primary BAG-trajectory model and re-estimating both terms (main-text Table 2). The specifications were: adding age × time; removing ICV; adding MRI field strength × time; adding baseline diagnosis × time; adding baseline ADAS-Cog-13 × time; and adding APOE ε4 allele dose × time. Covariates were entered as adjustments and were not crossed with sex. An age-only comparator was also included; it omitted education, ICV and their interactions with time from the primary model and therefore represents a separate comparator rather than a single-change sensitivity analysis.

#### Clinical progression

Clinical progression was also examined using Cox proportional-hazards models[21].

Baseline BAG, diagnosis, and covariates were taken at each participant's first retained MRI scan, and participants with dementia at that scan were excluded. Follow-up began at that scan and ended at the first subsequent ADNIMERGE clinical visit with a worse diagnosis than at baseline (CN to

MCI or dementia; MCI to dementia). Participants without progression were censored at their last clinical visit with an available diagnosis, and participants without follow-up time were excluded. The two transitions were modelled separately (MCI at baseline: n = 594, 216 events; CN at baseline: n = 487, 84 events).

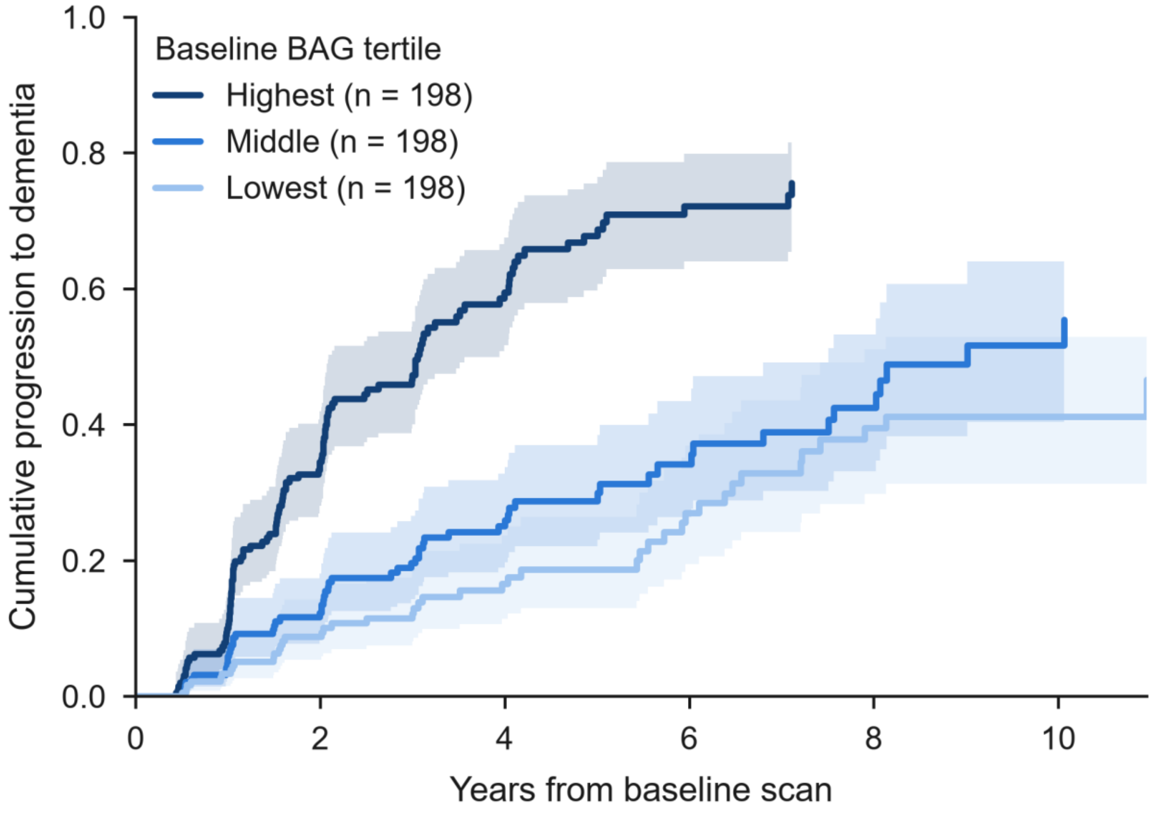


**Supplementary Figure S2.** Cumulative progression from mild cognitive impairment (MCI) to dementia by tertile of baseline brain age gap (BAG). Kaplan–Meier estimates are shown as one minus the estimated survival probability for 594 participants with MCI at their first retained MRI scan (198 per tertile; 216 progression events overall). Follow-up is measured in years from that scan. Shaded bands indicate 95% confidence intervals, calculated using Greenwood's variance on the log(-log) scale as described in the Supplementary Methods. Higher baseline BAG indicates an older-appearing brain relative to chronological age. The curves are unadjusted; adjusted associations are reported in the main text.

Models of BAG (per SD) adjusted for baseline age, education, ICV, sex, APOE ε4 carrier status, and baseline amyloid status. BAG × sex and BAG × amyloid interactions were tested by adding the corresponding product term. Models within amyloid strata (amyloid-positive; persistently amyloid-negative) omitted amyloid status and were fitted only when the stratum contained at least 15 events. Models with sex as the exposure omitted BAG and adjusted for baseline age, education, ICV, and APOE ε4 carrier status; hazard ratios compare men with women. Cox models were fitted with *statsmodels PHReg* using the Breslow method for tied event times. Cumulative progression from MCI to dementia by baseline-BAG tertile was estimated with the Kaplan–Meier method, with 95% confidence intervals from Greenwood's variance on the $log(-log)$ scale (Supplementary Figure S2).

#### Amyloid conversion

Among participants who were amyloid-negative at baseline, Cox proportional-hazards models were used to test whether baseline BAG predicted subsequent conversion to amyloid positivity. APOE ε4 was included as a positive control.

Follow-up began at the first retained MRI visit. The event was the first subsequent visit at which the participant was classified as amyloid-positive; participants who remained amyloid-negative were censored at their last visit with amyloid status available. Participants without follow-up after baseline were excluded, leaving 442 participants with 57 conversions. The model included baseline BAG (per SD), sex, APOE ε4 carrier status, baseline age, education, ICV, and the number of visits with amyloid status available, because participants with more assessments had more opportunity for a conversion to be observed. A BAG × sex interaction was tested in a separate model, and a model containing BAG alone was also fitted.

Amyloid status was determined from amyloid PET only (UC Berkeley amyloid PET classification in ADNI); CSF measures were not used to define amyloid status.

#### APOE ε4 allele dose

In BAG-trajectory models, APOE ε4 allele dose was coded with two indicator variables (one vs no ε4 allele; two vs no ε4 alleles), each entered with its interaction with time. The effects of one and two alleles on BAG level and rate of change were therefore estimated without assuming a linear allele-count trend, and the dose-response was tested with the contrast of two versus one allele. In survival models, APOE ε4 was coded as carrier status.

To assess how far the APOE ε4 associations were related to amyloid, APOE ε4 estimates were compared with and without adjustment for baseline amyloid status and its interaction with time. APOE ε4 effects within amyloid-negative and amyloid-positive participants were obtained as linear contrasts of a single APOE ε4 dose × amyloid × time model, and the APOE ε4 × amyloid terms tested whether these effects differed between strata. No formal mediation analysis was performed.
